# The evolving scenario of COVID-19 in hemodialysis patients

**DOI:** 10.1101/2022.06.09.22276185

**Authors:** Pasquale Esposito, Daniela Picciotto, Francesca Cappadona, Elisa Russo, Valeria Falqui, Novella Evelina Conti, Angelica Parodi, Laura Mallia, Sara Cavagnaro, Yuri Battaglia, Francesca Viazzi

## Abstract

**Background:** ‘Coronavirus disease 2019 (COVID-19) is a rapidly changing disease. So, in this study, we evaluated the evolution of COVID-19 presentation and course in hemodialysis patients (HD).

**Methods:** We retrospectively compared clinical data and outcomes of HD patients affected by COVID-19 during the first pandemic waves of 2020 (from March to December 2020-Group 1) with patients diagnosed with COVID-19 from September 2021 to February 2022 (Group 2), after the full completion of vaccination. Then, we distinguished among them patients responsive (antibody levels > 13 binding antibody units/ml) and unresponsive to the vaccine. We collected data on COVID-19 clinical presentation, laboratory examinations, and outcomes.

**Results:** Group 1 was constituted of 44 patients (69.3±14.6 years) and Group 2 of 55 patients (67.4±15.3 years). Among Group 2, fifty-two patients (95%) were vaccinated, 43 of them (83%) with three doses. Patients of Group 2, compared with Group 1, were more often asymptomatic (38 vs 10%, p=0.002), and reported less frequent fever and pulmonary involvement. At diagnosis, the Group 2 showed a significantly higher number of lymphocytes (0.97±0.45 vs 0.69±0.35 cells x109/L, p=0.008) and lower levels of circulating IL-6 (16±13.3 vs 41±39.4 pg/ml, p=0.002). Moreover, in Group 2, inflammatory parameters significantly improved after a few days from diagnosis. Patients of Group 2 presented a lower hospitalization rate (12.7 vs 38%, p=0.004), illness duration (18.8±7.7 vs 29.2±19.5 days, p=0.005), and mortality rate (5.4 vs 25%, p= 0.008). Finally, responders to the vaccination (80% of the vaccinated patients) compared with non-responders showed a reduction in infection duration and hospitalization (5 vs 40%, p=0.018).

**Conclusions:** COVID-19 presentation and course in HD patients have improved over time after the implementation of vaccine campaigns. However, due to the evolving nature of the disease, active surveillance is necessary.

## INTRODUCTION

Coronavirus disease 2019 (COVID-19) is characterized by a great heterogeneity of clinical presentation and outcomes, ranging from asymptomatic infections to severe forms leading to mortality. [1] So, many studies have evaluated the impact of different risk factors on the COVID-19 course and outcome.

These studies found that COVID-19-related mortality was associated with advanced age, male sex, and other conditions, including obesity, hypertension, cardiovascular disease, diabetes mellitus, chronic lung disease, and cancer. [2,3,4]

In this regard, patients developing kidney injury disease during COVID-19, or patients with pre-existing chronic kidney disease (CKD), are exposed to high mortality risk. [5] This consideration is also valid for patients undergoing maintenance hemodialysis (HD), who may be considered a distinct subgroup of COVID-19 patients. Indeed, these patients have some peculiarities that may influence disease course, including a high prevalence of comorbidities, frailty, a specific asset of the immune system. Moreover, logistical factors related to the organization of dialysis facilities may significantly impact dissemination and evolution of the infection in this setting. [6] These factors may also account for the high short-time mortality rate observed among HD patients with COVID-19 which, ranged between 20-30% of the affected patients especially in the surveys performed during the first phases of the pandemic.[7] Due to the high clinical impact, the COVID-19 pandemic has been the object of extensive studies investigating pathogenetic mechanisms, clinical management, diagnostic and therapeutic strategies. [8, 9] These efforts led to the rapid development of vaccination which has represented a game-change strategy showing high effectiveness in the prevention of severe disease, hospitalization, and death related to COVID-19. [10] This is a crucial point when considering HD patients because it is well-known that these patients present an immune dysfunction characterized by chronic systemic inflammation and premature aging of the immune system, which may impair their capacity to respond to the vaccines. [11,12] About COVID-19 vaccination, discordant results have been reported, indeed, although it seems that a high percentage of HD patients may mount humoral response after the vaccine, this response may be of short duration. [13,14] So, despite recent advances in fighting against COVID-19, HD patients still appear fragile and exposed to high risk. Moreover, apart from vaccination, other factors have emerged in the last months that may influence disease course, including the new virus variants and the availability of new therapeutic approaches, such as antiviral drugs and monoclonal antibodies. [15] Yet, there is no clear data on whether these new factors have affected the disease COVID-19 course in HD patients. Thus, this study aimed at investigating whether any difference exists in clinical presentation, time course, hospitalization, and mortality of COVID-19 patients undergoing dialysis between the first waves of the pandemic in 2020 and the last waves in late 2021-early 2022, after the full implementation of vaccination campaigns.

## PATIENTS AND METHODS

For this retrospective study we recruited three-weekly maintenance hemodialysis patients treated at San Martino, University Hospital of Genoa, Italy. We enrolled HD patients with COVID-19 confirmed by positive real-time reverse transcriptase (RT-PCR) assay for Severe Acute Respiratory Syndrome–CoronaVirus 2 (SARS-CoV-2) on nasopharyngeal swabs.

We compared patients who resulted positive for COVID-19 in two different periods: a) from March to December 2020 (Group 1), and b) from September 2021 to February 2022 (Group 2) after the full implementation of vaccination campaigns., i.e. after all patients were offered the possibility to receive at the least two doses of the COVID-19 vaccine on a voluntary basis.

So, within Group 2, we included both vaccinated patients and patients who voluntarily refused to get vaccinated. All the patients were vaccinated with COVID-19 mRNA BNT162b2 (Pfizer-BioNTech).

The study was performed according to the Declaration of Helsinki and was approved by the local Ethics Committee (N. Registro CER Liguria: 135/2020). All participants provided written informed consent before enrollment.

### Data collection

For each patient, we collected: i) demographics and clinical data, ii) clinical conditions, general laboratory, and chest X-ray findings at diagnosis, and iii) data on hospitalization, disease duration, and mortality. Laboratory examinations included: serum levels of inflammatory markers, such as high-sensitivity C-reactive protein (hs-CRP), procalcitonin (PCT), interleukin-6 (IL-6), ferritin, and lymphocyte count and neutrophil/lymphocyte ratio (N/L). IL-6 was determined at the central hospital laboratory by ELISA.

To assess the longitudinal evolution of COVID-19, the laboratory data were also evaluated at 7 and 14 days after the diagnosis.

In patients of Group 2, we also collected information on COVID-19 vaccination, including the number of doses administrated and serological response to the vaccination. The latter was assessed between the tenth and twentieth days after the second dose of the vaccine through the measurement of anti-S-RBD IgG levels, measured by Liaison SARS-CoV-2 trimeric S IgG Assay (Diasorin, Italy) and expressed as Binding Antibody Units (BAU)/m standardized according to the First WHO International Standard for anti-SARS-CoV-2 immunoglobulin. [16] Patients with antibody levels > 13 BAU/ml were considered responders to the vaccination, while patients with undetectable antibodies or levels <13 BAU/ml were considered non-responders.

### Statistical analysis

Data are presented as mean ± standard deviation (SD) or median with interquartile range (IQR), if not normally distributed (as evaluated by Shapiro Test). Student t-test, or nonparametric Mann-Whitney test, were used to assess the differences between patients affected by COVID-19 in different periods and between responders and non-responders. Mixed models for repeated measures, followed by Tukey’s multiple comparison tests were used to evaluate the temporal evolution of laboratory parameters in the different groups. Proportions for categorical variables were compared using Fisher’s exact test.

Time to event analyses were performed using: (i) Kaplan-Meier method for survival curves estimation and log-rank test to compare them; (ii) univariate Cox regression models: risk was reported as hazard ratios (HR) along with their 95% confidence intervals (CI).

Statistical calculations were performed by STATA package, version 14.2 (StataCorp, 4905 Lakeway Drive, College Station, Texas 77845 USA). The null hypothesis was rejected for values of P < 0.05.

## RESULTS

### Patient characteristics

Group 1 was constituted of 44 HD patients (69.3±14.6 years, 14 males) with a length of time on dialysis of 67±58.4 months. Thirty-four patients (77%) were hypertensive, 12 (27.3%) diabetic and 30 (68.2%) had a history of cardiovascular disease, while four patients (9%) had a history of transplantation (3 kidney, 1 liver).

Group 2 was constituted of 55 HD patients (67.4±15.3 years, 39 males) with a dialysis vintage of 39±29 months. Forty-eight patients (87.3%) were hypertensive, 23 (41.8%) diabetic and 22 (40%) had a history of cardiovascular disease, while eight patients (14%) had a history of transplantation (7 kidney, 1 liver)

Overall, six patients (11%) in the Group 2 presented a previous history of COVID-19 infection, two of them had the first COVID-19 diagnosis in 2020 and then were included also in Group 1, while the remaining four had the first infection in 2021.

The comparison between the groups showed that patients in the Group 2 presented a significantly lower length of time on dialysis, a higher prevalence of diabetes, and a lower prevalence of cardiovascular disease (Table 1).

**Table 1.** Demographics and medical history at admission of HD patients affected by COVID-19 during the different periods evaluated.

|  | **Group 1** | **Group 2** | **p** |
| --- | --- | --- | --- |
| **N,** | 44 | 55 |  |
| **Age, yr** | 69.3±14.6 | 67.4±15.3 | 0.6 |
| **Men, n (%)** | 24 (54.5) | 39 (70.9) | 0.09 |
| **BMI, kg/m<sup>2</sup>** | 24.1±4.9 | 25±5.2 | 0.4 |
| **Length of time on dialysis, months** | 67±58.4 | 39±29 | 0.015 |
| **Comorbidities** |  |  |  |
| **Hypertension, n (%)** | 34 (77) | 48 (87.2) | 0.3 |
| **Diabetes mellitus, n (%)** | 12 (27.3) | 23 (41.8) | 0.03 |
| **CVD, n (%)** | 30 (68.2) | 22 (40) | 0.008 |
| **Autoimmune disorders, n (%)** | 1 (2) | 5 (9) | 0.2 |
| **Prior transplant, n (%)** | 4 (9) | 8 (14.5) | 0.5 |
| **History of COVID-19, n (%)** | - | 6 (11) | - |
| **Vaccinated patients, n (%)** | - | 52 (94.5) | - |
| **Vaccinated with booster dose, n (%)** | - | 43 (78.2) | - |
| **Responders after the second dose of vaccine, n (%) \*** | - | 39 (80) |  |

Abbreviations: Coronavirus disease-19, COVID-19; hemodialysis, HD; body mass index, BMI; cardiovascular disease, CVD.
Group 1, from March to December 2020; Group 2, from September 2021 to February 2022
\* data available for 49 patients

Among patients of Group 2, fifty-two (95%) were vaccinated and 43 of them (83%) had received the third booster dose of vaccination at the time of COVID-19 diagnosis. A percentage that was in line with that observed in the whole HD patient population at our Center (where, as of February 2022, 265/282 patients-i.e. 94%, were vaccinated).

The mean interval from COVID-19 diagnosis and last vaccination dose was 112±58 days. According to the serum levels of anti-S-RBD IgG available in 49 vaccinated patients of Group 2, 39 patients (80%) were responsive to the second dose of the vaccine.

At the disease presentation, non-responders had a significant lower BMI than responders, and reported a high prevalence of history of autoimmune disorders (S1 Table).

### Clinical presentation and radiological findings

In Group 2, there was a significantly higher number of asymptomatic patients when compared with patients in Group 1 (38% vs 10%, p=0.002) (Table 2).

**Table 2.** Clinical presentation and basal radiological findings of HD patients affected by COVID-19 during the different periods evaluated.

| COVID-19 presentation<br>n (%) | Group 1<br>(n, 44) | Group 2<br>(n, 55) | p |
| --- | --- | --- | --- |
| **Patients symptomatic at diagnosis** | 40 (90) | 35 (62) | 0.002 |
| **Fever** | 28 (63.3) | 17 (31) | 0.002 |
| **Cough** | 17 (38.6) | 17 (31) | 0.5 |
| **Dyspnea (exertional or rest)** | 14 (32) | 4 (7) | 0.003 |
| **Dysgeusia/anosmia** | 10 (23) | 2 (3.6) | 0.005 |
| **Fatigue/malaise** | 3 (6.8) | 1 (1.8) | 0.8 |
| **Gastrointestinal** | 5 (11.4) | 1 (1.8) | 0.08 |
| **Myalgia** | 6 (13.6) | 0 | 0.006 |
| **SatO<sub>2</sub>, %** | $0.95\pm 0.03$ | $0.96\pm 0.03$ | 0.16 |
| **SBP, mmHg** | 124.7±28 | 138±25 | 0.018 |
| **DBP, mmHg** | 62.7±13 | 67.7±14 | 0.17 |
| **Chest X-ray (clinical indication), n (%)** | 17 (38.6) | 6 (10.9) | 0.001 |
| **Lung consolidations** | 14 (82) | 5 (83) |  |
| **Pleural effusion** | 2 (4.5) | 2 (3.6) |  |

Abbreviations: Coronavirus disease-19, COVID-19; hemodialysis, HD; Arterial blood oxygen saturation, SatO<sub>2</sub>; Diastolic blood pressure, DBP; Systolic blood pressure, SBP.
Group 1, from March to December 2020; Group 2, from September 2021 to February 2022

Initial symptoms were various, being fever, cough, and shortness of breath the most common ones.

However, patients in the Group 2 presented a lower incidence of fever, such as dysgeusia, diarrhea, and myalgia. Oxygen saturation and diastolic blood pressure were not different between the two groups, whereas systolic blood pressure was significantly higher in Group 2 (138±25 vs 124.7±28 mmHg, p=0.018). Basal chest X-ray was requested on clinical indication in 17 patients (38%) of Group 1 and in 6 patients (10.9%) of Group 2 (p= 0.001), mostly showing lung consolidations.

### Basal laboratory characteristics

The comparison of biochemical parameters collected at diagnosis showed that patients affected by COVID-19 in late 2021, when compared with patients of the 2020 pandemic waves, presented a significantly higher number of lymphocytes (0.97±0.45 vs 0.69±0.35 cells x109/L, p=0.008), and lower values of N/L [4.2 (3.5) vs 5.8 (5.4), p=0.009] and LDH (220±173 vs 254±98 U/L, p=0.003) (Figure 1). Considering the inflammatory markers, patients in the Group 2 had comparable levels of PCTI and hs-CRP but significantly lower levels of circulating IL-6 (16±13.3 vs 41±39.4 pg/ml, p=0.002) and ferritin [242 (407) vs 552.5 (903) 𝜇g/L, p=0.021] (Table 3).

**Figure 1.**
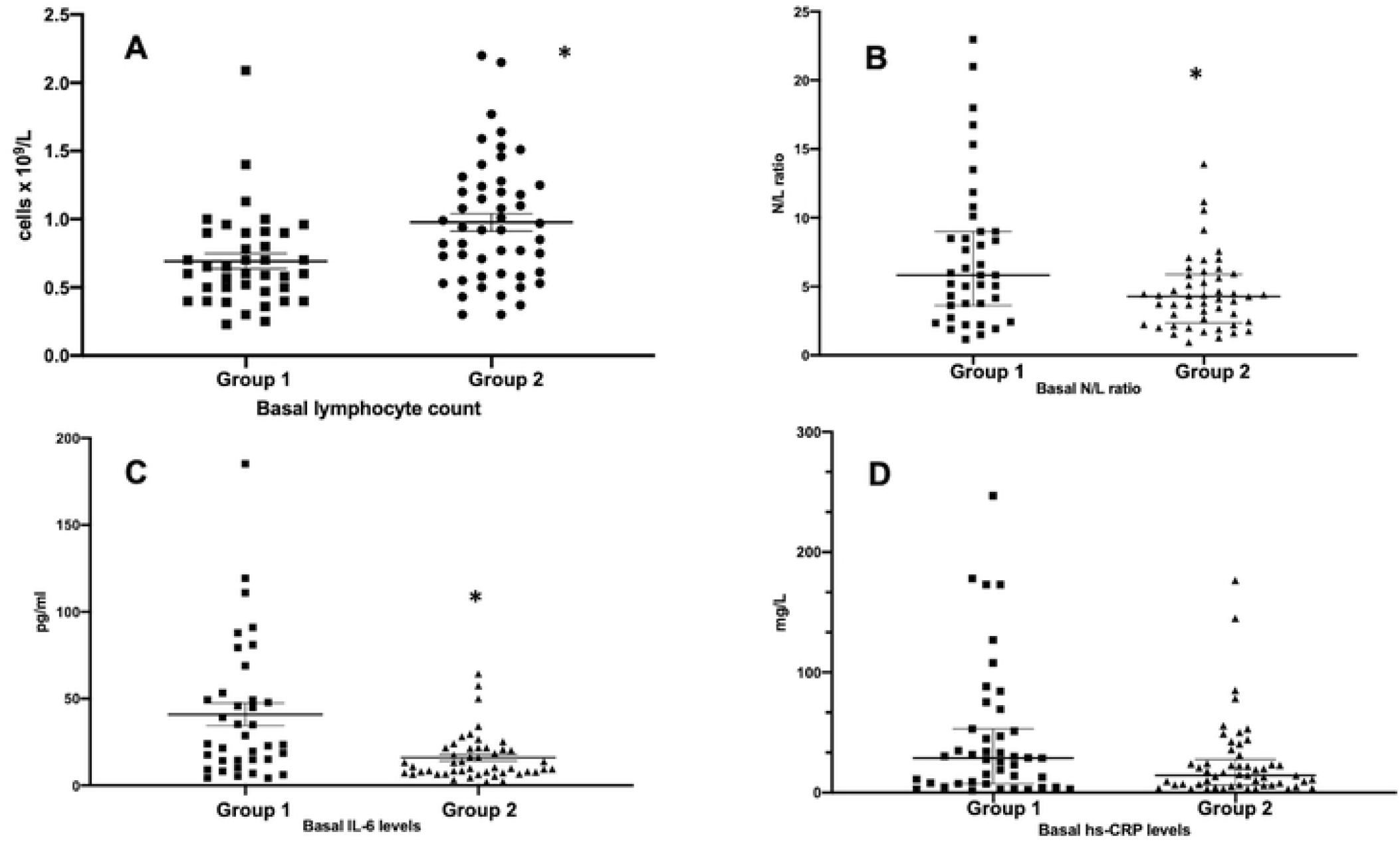
Comparisons of inflammatory parameters between HD patients affected by COVID-19 during the first pandemic waves of 2020 and last wave of 2021-early 2022. Data are expressed as mean with standard error in Figure 1A and 1C and median with interquartile range in Figure 1B and 1D. * p<0.01 Group 1, from March to December 2020; Group 2, from September 2021 to February 2022

**Table 3.** Basal laboratory parameters of HD patients affected by COVID-19 during the different periods evaluated.

|  | **Group 1** | **Group 2** | **p** |
| --- | --- | --- | --- |
| **N,** | 44 | 55 |  |
| **WBC Count, ×10<sup>9</sup>/L** | 5.5±3.2 | 5.5±1.8 | 0.2 |
| **Neutrophil count, ×10<sup>9</sup>/L** | 4.3±3.2 | 3.6±1.4 | 0.5 |
| **Neutrophils, (percent of WBC)** | 72.2±17.7 | 66.7±10.9 | 0.003 |
| **Lymphocyte count, × 10<sup>9</sup>/L** | 0.69±0.35 | 0.97±0.45 | 0.008 |
| **Lymphocytes, (percent of WBC)** | 14.7±8.9 | 18.2±8.1 | 0.024 |
| **N/L ratio** | 5.8 (5.4) | 4.2 (3.5) | 0.009 |
| **Platelet Count, ×10<sup>9</sup>/L** | 200±127 | 194±70 | 0.3 |
| **Hemoglobin, mmol/L** | 6.7±0.99 | 6.95±0.8 | 0.19 |
| **Albumin, g/L** | 34±4 | 36±7 | 0.07 |
| **LDH, U/L** | 254±98 | 220±173 | 0.003 |
| **PCT, µg/L** | 1.81 (3.3) | 1.40 (1.5) | 0.25 |
| **hs-CRP, mg/L** | 28.8 (45.6) | 14.4 (16.5) | 0.08 |
| **IL-6, pg/ml** | 41±39.4 | 16±13.3 | 0.002 |
| **Ferritin, µg/L** | 552.5 (903) | 242 (407) | 0.02 |

Data are expressed as mean±SD or median (interquartile range)
Abbreviations: Coronavirus disease-19, COVID-19; hemodialysis, HD; white blood cells, WBC; neutrophil/lymphocyte ratio, N/L; lactate dehydrogenase, LDH; interleukin-6, IL-6; Procalcitonin, PCT; high-sensitivity C-reactive protein, hs-CRP.
Group 1, from March to December 2020; Group 2, from September 2021 to February 2022

Patients responsive to the vaccination compared with non-responders did not show significant differences in basal laboratory parameters (S2 Table).

### Time evolution of inflammatory parameters

Evaluating the temporal course of the principal laboratory parameters, we found that in Group 1, no significant changes in inflammatory parameters occurred over time. On the opposite, in Group 2, we observed a rapid improvement of some parameters, such as white blood cell, neutrophil, and lymphocyte count that significantly increased one week after the diagnosis, accompanied by a significant reduction of hs-CRP levels [14.4 (16.5) at diagnosis vs 7.4 (20) mg/L at day 7, p=0.002] (S3 Table).

### Clinical outcomes

As for the Group 1, 16 patients (38%) were admitted to the hospital because of worsening clinical conditions, while the remaining patients underwent outpatient hemodialysis.

No antiviral treatment was prescribed, while 17 patients (63%) received treatment with hydroxychloroquine. Overall, 11 patients died (25%) after a mean of 13±10.5 days from COVID-19 diagnosis, whereas 33 patients resulted negative for SARS-CoV2 after 29.2±19.5 days from diagnosis. Among Group 2, 7 patients (12.7%) were admitted to the hospital. Eight patients (14.5%) were treated with anti-COVID-19 monoclonal antibodies (7 with the combination casirivimab+ imdevimab and 1 with bamlanivimab + etesevimab), five patients received steroids, and one patient anakinra, an interleukin-1 receptor antagonist. None of the patients in the Group 2 was treated with hydroxychloroquine. Overall, 3 patients died (5.4%) after a mean of 7.3±1.1 days from COVID-19 diagnosis, whereas the remaining 52 patients resulted negative for SARS-COV2 after 18.8±7.7 days from diagnosis.

The comparison of clinical outcomes showed that Group 2 presented significantly lower mortality, as well as lower hospitalization and infection duration (Table 4, Figure 2).

**Table 4.** Clinical outcomes of HD patients affected by COVID-19 during the different periods evaluated.

|  | **Group 1** | **Group 2** | **p** |
| --- | --- | --- | --- |
| **N,** | 44 | 55 |  |
| **Illness duration, days\*** | 29.2±19.5 | 18.8±7.7 | 0.005 |
| **Hospitalized patients, n (%)** | 17 (38) | 7 (12.7) | 0.004 |
| **Deaths, n (%)** | 11 (25) | 3 (5.4) | 0.008 |
| **Time from diagnosis to death, day** | 13±10.5 | 7.3±1.1 | 0.4 |

\*Illness duration was calculated only in survivors from the date of the first positive RT-PCR assay for SARS-COV2 infection to the date of the two consecutive negative RT-PCR assays.

**Figure 2.**
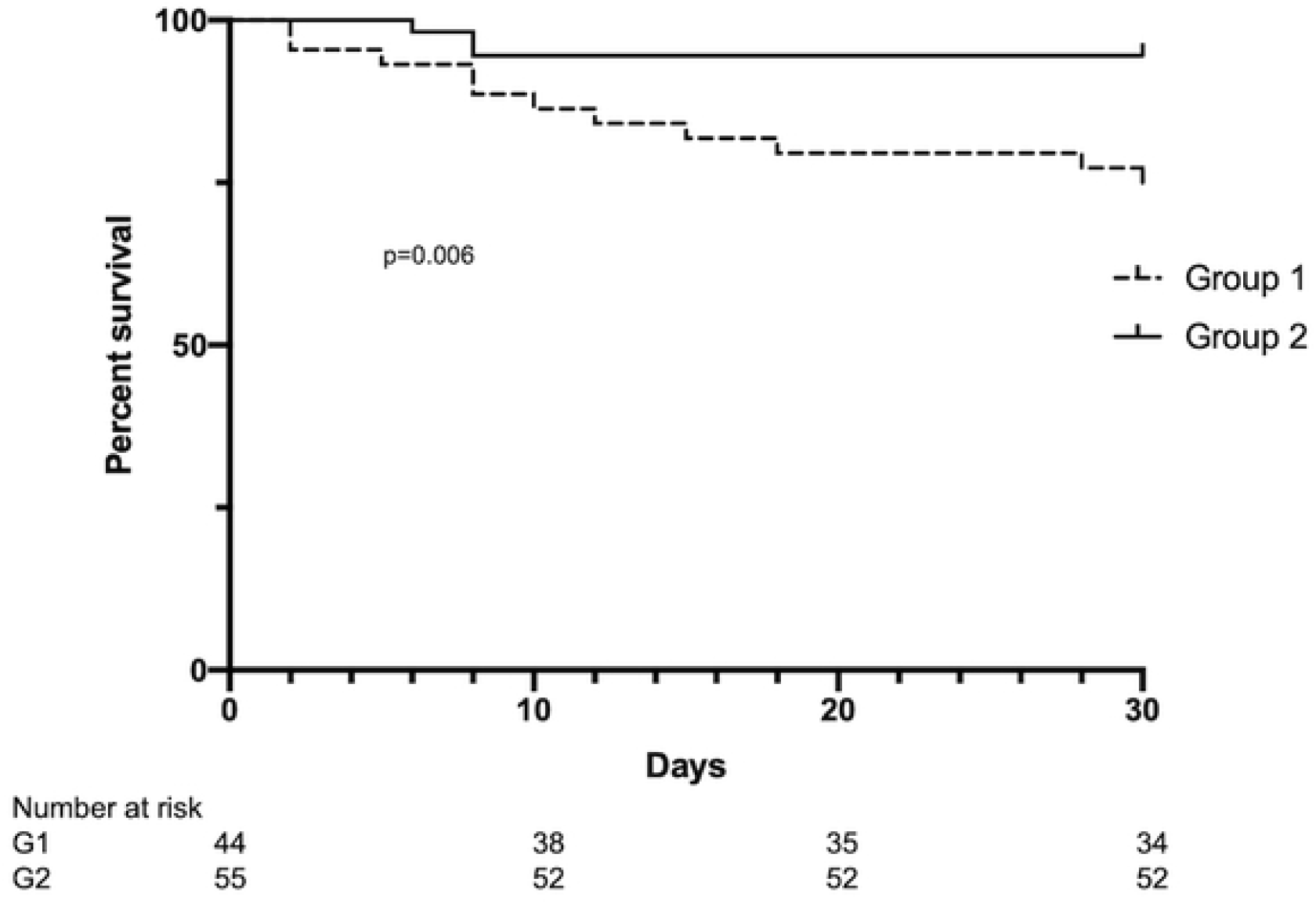
Kaplan-Meier curves of 30-day survival in HD patients affected by COVID-19 during the first pandemic waves and the last wave of 2021. Group 1, from March to December 2020; Group 2, from September 2021 to February 2022

Then, we performed a univariate Cox analysis in the entire cohort of both the groups of patients, considering age, gender, length of time on HD, CVD, diabetes, and vaccine as variables. We observed that history of CVD was positively associated with the mortality (with HR 1.18, CI 1.005-1.39, p=0.043), whereas vaccination was negatively (HR 0.26, CI 0.07-0.96, p=0.04). Multivariate analysis was not performed due to the low number of events. Finally, comparing responders to the vaccination with non-responders, we observed that responders presented a significant reduction in infection duration (17.6±6.4 vs 23.9±7.9 days, p=0.02) and hospitalization rate 5% vs 40%, p=0.018), while mortality was not different between the two groups (S4 Table).

## DISCUSSION

In this study, we evaluated the temporal evolution of the COVID-19 pandemic in HD patients, comparing patients affected by COVID-19 during the first pandemic waves of 2020 with patients diagnosed with COVID-19 in late 2021-early 2022, after each patient has had access to at least two doses of the COVID-19 vaccine.

We selected these periods to have the opportunity to assess the clinical impact of full vaccination campaigns and the introduction of specific antiviral treatments developed and diffused during 2021. We observed a significant change in the COVID-19 course in HD and, in particular, during the second period, the patients presented a milder form of the disease and better clinical outcomes.

These patients, compared with those of the first pandemic waves, were more often asymptomatic and reported less frequent fever and specific symptoms, like myalgia and asthenia. Moreover, they presented higher blood pressure values and less COVID-19-related pulmonary involvement. Significant differences were also found in laboratory examinations that showed in patients of Group 2 a minor degree of lymphopenia and reduced levels of LDH and inflammatory parameters, including ferritin and IL-6 levels.

These findings are of particular importance because in COVID-19 patients the alterations of these parameters have been associated with the disease severity and mortality risk.[17] Moreover, the time course of laboratory examinations was also different between the two groups of patients, with patients of Group 2 showing a significant reduction of inflammatory parameters after a few days from diagnosis.

The better clinical and laboratory profile of patients affected by COVID-19 in late 2021-early 2022 corresponded to a significant improvement of the clinical outcomes, evaluated as the duration of the infection, hospitalization rate, and mortality. Then, while the mortality rate observed during the first phases of the pandemic was in line with that reported in previous studies on HD patients (25%), it decreased in patients affected by COVID-19 in the late phases (5.4%).

Overall, our data show that, at least in maintenance HD patients, COVID-19 has evolved, and disease characteristics and short-term sequelae are changed.

It is conceivable that there are many reasons for these changes. First, we noticed some differences in the patient characteristics, with patients of the first waves presenting a significantly higher length of time on dialysis, a higher prevalence of cardiovascular disease, and a lower prevalence of diabetes, as compared with the patients of the Group 2. Although we found only a weak relationship between a history of CVD and death in our cohort, we cannot rule out the possibility that underlying chronic disease may have conditioned COVID-19 expression. [18]

It is plausible that a relevant role in the change of COVID-19 presentation and evolution has been played by the vaccination. In this regard, we were favored by the active vaccine strategy promoted by the Italian healthcare system which led to the high percentage of vaccinated patients present in our cohort. This condition allowed us to observe that, despite the initial concerns, also in HD patients, the vaccination was associated with a reduction in mortality and a significant improvement in clinical outcomes. [19] Moreover, we noticed an elevated percentage of patients mounting a sufficient immune response to COVID-19 vaccination, which conceivably further increased after administration of the third booster dose. [20,21] We found that compared to responsive subjects, the non-responsive ones presented higher hospitalization and disease duration, while, possibly due to the low sample size, the mortality rate was not different. These data reinforce the idea that vaccination is a game-change strategy in protecting from COVID-19 severe complications [22]. Moreover, they suggest that within vaccinated HD patients, a partial heterogeneity of COVID-19 presentation and course may persist, probably driven by a different underlying immune status.

Apart from vaccination, an important aspect to consider when evaluating COVID-19 pandemic evolution is the impact of the new SARS-CoV2 variants.

It is well-known that SARS-CoV2 is a mutating virus, and the emergence of the variants of concern may affect the disease course because they have different transmissibility and pathogenicity.[23]

Specifically, in Italy, in the first phases of the pandemic of 2020, the predominant variants were the original Wuhan strain SARS-CoV-2 and B.1.1.7 (Alpha), while in late 2021, B.1.617.2 (Delta) and B.1.1.529 (Omicron) variants were the most prevalent ones. [24,25] The prevalence of different virus variants may explain some differences we found in our cohorts. For example, the Omicron variant replicates faster than all other SARS-CoV-2 variants and has been associated with less dysgeusia and pneumonia. [26,27] Finally, an additional factor contributing to the change of COVID-19 course in HD patients is the availability of virus-specific treatments, such as monoclonal antibodies or antiviral drugs. These treatments, introduced in 2021, may represent an opportunity to reduce the risk of COVID-19–related hospitalization or death.[28] However, these new therapeutic strategies are still limited in HD patients, in whom, for instance, the new oral antiviral drugs are contraindicated.[29] Moreover, considering the use of monoclonal antibodies, we found that in our patients infected during the late pandemic wave, the most used antiviral treatment was the combination of casirivimab+ imdevimab, which is poorly effective against the Omicron variant.[30] The choice of this treatment was influenced by the fact that at the beginning of 2022, Italy’s health system suffered from a shortage of sotrovimab, a monoclonal antibody active against the Omicron variant.[31] So, it seems that the potentialities of COVID-19 treatment have not been fully exploited until now, while the development of antiviral treatments prescribable also for HD patients seems mandatory.

We are aware that the relatively small cohort of studied patients, together with the inability to discriminate and quantify the factors influencing the disease course, including the lack of data on virus variants, are the main weaknesses of this study. Moreover, we have no data on the immune response to the third booster dose of the vaccine that could have reinforced the specific immune response against the infection. To overcome these limitations, there is the need for large prospective longitudinal studies evaluating the time course of both humoral and cellular response to the vaccination and molecular studies on virus variants’ epidemiology and their clinical impact.

## CONCLUSIONS

Our data show that COVID-19 presentation and course in HD patients have improved over time, especially after the implementation of vaccine campaigns, even if this change was probably a result of the cooperation of different factors rather than the effect of a single element.

However, since COVID-19 is a continuing evolving disease, many questions, such as the duration of immunity, the effectiveness of the vaccinations, the need for more vaccine doses, the effects of antiviral drugs against new virus variants, and long-term outcomes are still open. [32] So, while continuing the promotion of vaccination and the development of innovative therapeutic strategies remain of paramount importance, active surveillance of the COVID-19 course should be warranted.

## Data Availability

All relevant data are within the manuscript and its Supporting Information files.

## Acknowledgements

The authors thank the Dialysis Units of IRCCS Policlinico San Martino of Genova.

## Supporting information

S1 Table. Demographics and medical history at admission of HD patients affected by COVID-19 according to the response to the second dose of COVID-19 mRNA vaccine.

S2 Table. Basal laboratory parameters of HD patients affected by COVID-19 according to the response to the second dose of COVID-19 mRNA vaccine.

S3 Table. Time course of laboratory parameters in HD patients affected by COVID-19 during different periods.

S4 Table. Clinical outcomes of HD patients affected by COVID-19 according to the response to the second dose of COVID-19 mRNA vaccine.

## Notes

### Competing Interest Statement

The authors have declared no competing interest.

### Funding Statement

The funders had no role in study design, data collection and analysis, decision to publish, or preparation of the manuscript.

